# Optimal cutoffs for the Montreal Cognitive Assessment among English and Spanish speaking Latinos

**DOI:** 10.64898/2026.01.26.26344881

**Authors:** Jaime Perales-Puchalt, Andrew J. Aschenbrenner, Maria Marquine, Katya Rascovsky, Adam Parks

## Abstract

The Montreal Cognitive Assessment (MoCA) is widely used to screen for cognitive impairment, yet commonly applied cutoff scores have been found to perform poorly among US Latinos. Prior studies relied on small samples, combining persons with mild cognitive impairment (MCI) and dementia into a single group, or failing to account for multiple intersecting demographic factors. We identified optimal MoCA cutoffs for MCI and dementia among US Latinos while addressing these limitations. We analyzed cross-sectional data from the National Alzheimer’s Coordinating Center Unified Data Set. Participants included English- and Spanish-speaking Latinos who completed testing in their primary language. Research diagnostic groups consisted of cognitively unimpaired (CU), MCI, and dementia. Groups were further stratified by testing language, age, and level of education. Diagnostic accuracy and receiver operating characteristic (ROC) analyses were performed. The Youden Index was used to determine the optimal cutoff score. Of the 1,673 participants in the total sample, 46% completed the MoCA in Spanish and 54% in English, 54% were CU, and 28% had MCI and 19% had dementia. Area under the curve (AUC) values for CU vs. MCI were 0.70 for Spanish-tested participants and 0.79 for English-tested participants, while values for MCI vs. dementia were 0.85 and 0.89 for the Spanish and English tested participants, respectively. Overall AUC values were 0.76 for CU vs. MCI and 0.86 for mild cognitive impairment vs. dementia. Optimal cutoffs were consistently found to be lower among participants tested in Spanish, those older than age 75, and participants with the fewest years of education, with some optimal cutoffs shown to be substantially lower than the traditionally used standard cutoff. These findings provide cutoffs that better reflect differences amongst language and demographic groups. We also provide a scoring calculator for clinical and research use.

## Introduction

Dementia is a leading cause of mortality in the US that causes substantial disability [1], and imposes major costs on the healthcare system [2]. Reducing dementia-related health disparities among groups at disproportionate risk is a national priority [3, 4]. Latinos face a higher risk of dementia compared to non-Latino Whites [5]. The number of Latinos with dementia is projected to increase by more than 800% between 2012 to 2060, impacting an estimated 3.5 million individuals [6].

Besides dementia risk disparities, Latinos with dementia and mild cognitive impairment (MCI) are disproportionately undiagnosed or diagnosed late or inaccurately, compared to non-Latino Whites [7-9]. Disparities in diagnosis may reduce Latinos’ chances of receiving optimal care or increase unnecessary distress [10, 11]. This suboptimal care likely contributes to Latinos experiencing a disproportionate prevalence of behavioral and psychological symptoms [12, 13], and informal dementia caregiver depression compared to non-Latino Whites [14].

Cognitive screening is a crucial step to the diagnosis of dementia and MCI in dementia care guidelines [15]. Cognitive screening tools are especially important in busy primary care clinics where there are already time constraints that can cause unnecessary stress to patients and families [16]. Despite this need, few cognitive screening tools have been validated for use with US Latinos, including the Mini Mental State Exam (MMSE), Telephone Interview for Cognitive Status (TICS), Dementia Questionnaire, and Montreal Cognitive Assessment (MoCA) [17-21].

The MoCA is the second most widely used worldwide is the and has demonstrated greater sensitivity than the MMSE for detecting MCI [22]. Prior studies assessing the diagnostic performance of the MoCA among Latinos in the US contain several limitations, including inadequate sample sizes, failure to distinguish MCI from dementia, not discern between those tested in their primary or secondary language (for which proficiency level might vary and can confound performance with secondary language skills), and not stratify results by relevant factors in Latino cognitive research that may impact cognitive function and testing-taking skills such as age, education, language [18-20, 23].

In the current manuscript, we aim to identify optimal cutoffs for MCI and dementia among US Latinos. We address limitations of the extant research by using a larger sample, separating MCI and dementia reference standards, for restricting analyses to individuals tested in their primary language, and stratifying results by key demographic factors. We hypothesize that the diagnostic accuracy tests will show that lower cutoff scores on the MoCA are more accurate in discriminating MCI and dementia for Spanish tested participants, older adults (Age > 75 years), and individuals with lower educational attainment compared to English speakers, younger participants, and those with higher educational attainment. We also provide a spreadsheet to calculate the most appropriate MoCA cutoff depending on an individual’s language, age, and educational level (see Supplemental Material).

## Methods

### Participants

Data for participants were drawn from the National Alzheimer’s Disease Coordinating Center (NACC) database (2015-June 2024), which contains the Uniform Data Set (UDS), a standardized collection of demographic, clinical, and cognitive data from all National Institute on Aging funded Alzheimer Disease Research Centers in the United States [24, 25]. For the current analyses, we included participants aged 55 years or older who self-identified as Latino and completed the MoCA. Only the first assessment at which a participant completed the MoCA was included. To ensure participants were tested in their primary language, their primary language (English or Spanish), was required to match the language of MoCA administration.

### Measures

Primary variables of interest included the MoCA total uncorrected raw score, clinician-determined diagnosis of cognitive status at the time of assessment (normal cognition, MCI, or dementia), self-rated years of education, age at assessment, and language of MoCA administration (Spanish or English).

### Data Analysis

All statistical analyses were conducted in R version 4.5.1. We used the pROC [26] and cutpointr [27] packages to estimate sensitivity, specificity, positive predictive value (PPV), negative predictive value (NPV), and area under the curve (AUC) for MoCA cutoff scores distinguishing a) cognitively unimpaired participants (CU) from those with MCI and b) participants with MCI from those with dementia. Optimal cutoff scores were defined as the values that maximized the Youden Index. Analyses were stratified by language of testing, English or Spanish, age group, younger than 75 years versus 75 years and older, and educational attainment, categorized as fewer than 6 years, 7 to 11 years, 12 to 15 years, and 16 or more years, and 95% confidence intervals for AUC values were generated using bootstrap sampling.

## Results

The final analytic sample included 1,673 Latino participants. Table 1 shows the characteristics of the sample: 65% (n=1,081) women, 54% (n=899) CU, 28% (n=461) MCI, and 19% (n=313) dementia. Age ranged from 55 to 96 and averaged 71 (SD=8). Years of education ranged from 0 to 20 years and averaged 13.2 (SD=4.5) years. The uncorrected MoCA Total Raw Score ranged from 0 to 30 with a mean of 21 (SD=6). The MoCA Total Score corrected for education (one additional point given to those with less than or equal to 12 years of education ranged from 1 to 30 with an average score of 21 (SD=6). Participants tested in Spanish were older, more often women, had fewer years of education, were more likely to be diagnosed with MCI or dementia, and had lower MoCA Total Raw and corrected scores compared to those tested in English.

**Table 1.**
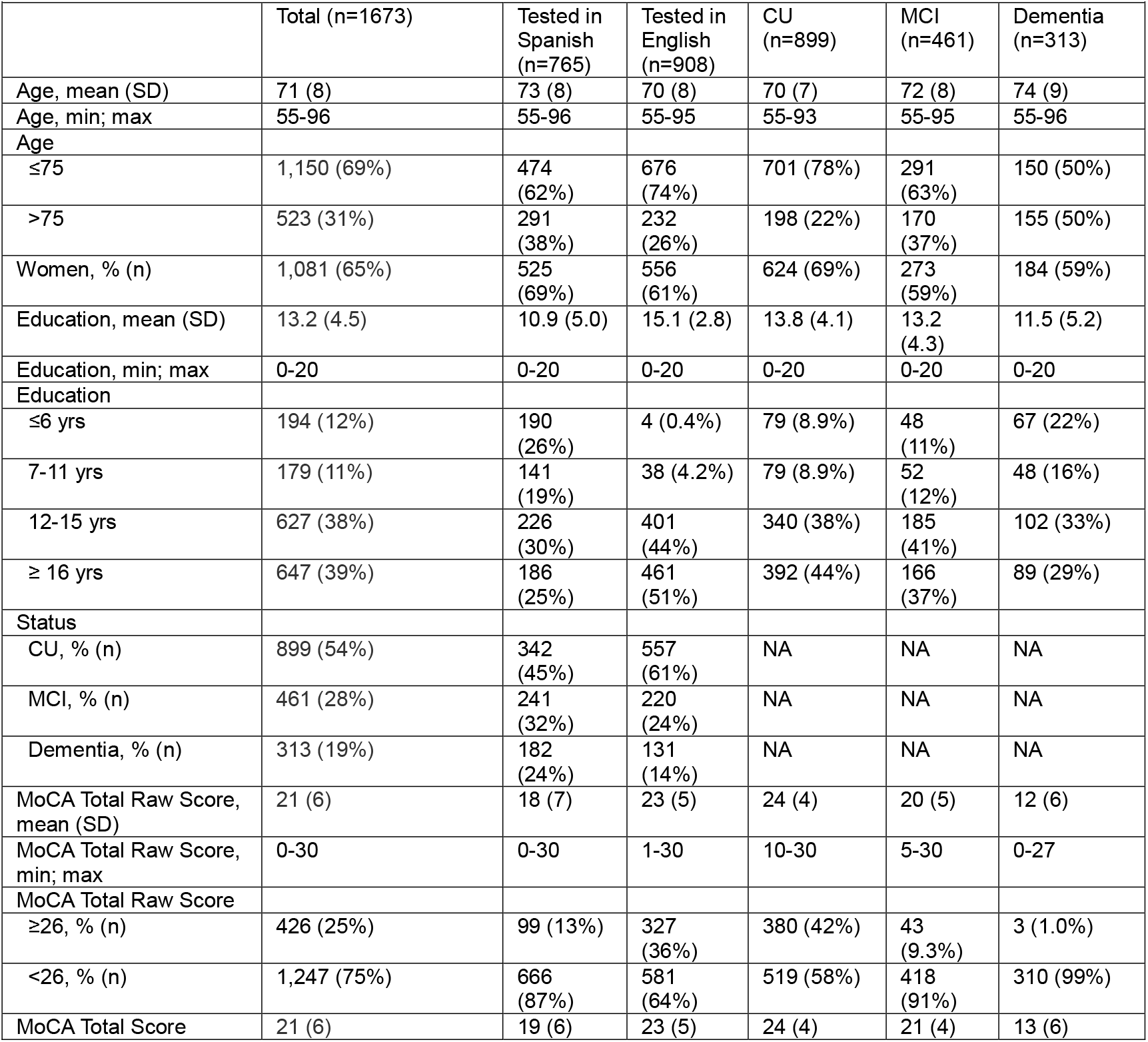
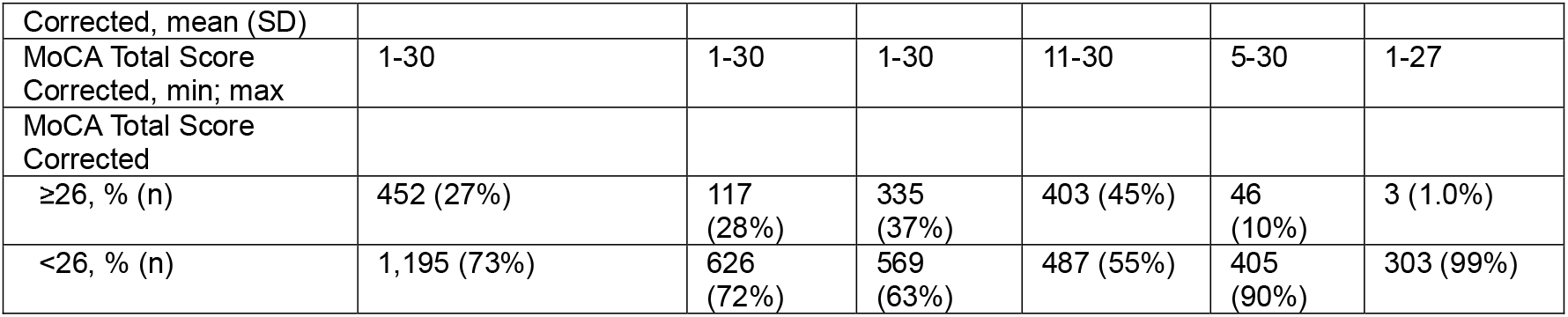
Characteristics of the sample.

Table 2 shows the diagnostic performance of the MoCA for detecting MCI or dementia using the original cutoff score of 26, based on both raw and education corrected scores. Using the MoCA Total Raw Score, sensitivity was 0.94, specificity was 0.42, the positive predictive value was 0.58, and the negative predictive value was 0.89. Sensitivity was slightly higher and specificity lower among those tested in Spanish compared to those tested in English, regardless of whether the MoCA raw or education corrected scores were used. For example, using the MoCA Total Raw Score, the sensitivity was 0.96 versus 0.92 and specificity was 0.23 versus 0.54 for those tested in Spanish compared to English tested participants. Sensitivity and specificity did not meaningfully differ between raw and education corrected scoring.

**Table 2.** Diagnostic performance of the MoCA in detecting impairment using the current cutoff of 26 for MCI and 18 for dementia.

|  |  |  |  |  |  |  |  |  |  |
| --- | --- | --- | --- | --- | --- | --- | --- | --- | --- |
|  |  | Normal cognition vs. MCI |  |  |  | MCI vs. Dementia |  |  |  |
|  |  | Sensitivity | Specificity | PPV | NPV | Sensitivity | Specificity | PPV | NPV |
| MoCA Total Raw Score | Total | 0.91 | 0.42 | 0.45 | 0.90 | 0.82 | 0.74 | 0.68 | 0.86 |
|  | Tested in Spanish | 0.93 | 0.23 | 0.46 | 0.82 | 0.87 | 0.61 | 0.63 | 0.87 |
|  | Tested in English | 0.88 | 0.54 | 0.43 | 0.92 | 0.74 | 0.88 | 0.78 | 0.85 |
| MoCA Total Score Corrected | Total | 0.90 | 0.45 | 0.45 | 0.90 | 0.79 | 0.78 | 0.71 | 0.85 |
|  | Tested in Spanish | 0.92 | 0.29 | 0.47 | 0.84 | 0.85 | 0.66 | 0.65 | 0.85 |
|  | Tested in English | 0.87 | 0.55 | 0.44 | 0.92 | 0.72 | 0.90 | 0.82 | 0.84 |

Tables 3 and 4 show the AUC values and Youden Index for individual MoCA scores across subsamples. Discrimination was stronger for dementia than for mild cognitive impairment. For example, AUC values in the total sample were 0.86 for dementia and 0.76 for MCI. Optimal cutoff scores were consistently higher among participants tested in English compared to those tested in Spanish, including cutoffs of 25 versus 23 for mild cognitive impairment and 17 versus 16 for dementia.

**Table 3.** Optimal MoCA cutoffs to distinguish between normal cognition and MCI.

|  |  |  |  |  |  |  |  |  |  |
| --- | --- | --- | --- | --- | --- | --- | --- | --- | --- |
| Score | Sensitivity |  |  | Specificity |  |  | Youden's Index |  |  |
|  | Total | Spanish | English | Total | Spanish | English | Total | Spanish | English |
| 20 | 0.39 | 0.52 | 0.25 | 0.86 | 0.73 | 0.95 | 0.25 | 0.25 | 0.2 |
| 21 | 0.46 | 0.59 | 0.34 | 0.82 | 0.66 | 0.91 | 0.28 | 0.25 | 0.25 |
| 22 | 0.57 | 0.68 | 0.45 | 0.77 | 0.60 | 0.88 | 0.34 | 0.28 | 0.33 |
| 23 | 0.66 | 0.76 | 0.55 | 0.73 | 0.54 | 0.84 | **0.39** | **0.3** | 0.39 |
| 24 | 0.74 | 0.80 | 0.67 | 0.64 | 0.46 | 0.75 | 0.38 | 0.26 | 0.42 |
| 25 | 0.84 | 0.88 | 0.80 | 0.54 | 0.34 | 0.66 | 0.38 | 0.22 | **0.46** |
| 26 | 0.91 | 0.93 | 0.88 | 0.42 | 0.23 | 0.54 | 0.33 | 0.16 | 0.42 |
| 27 | 0.96 | 0.98 | 0.93 | 0.32 | 0.17 | 0.41 | 0.28 | 0.15 | 0.34 |

AUC: Total = 0.76, CI = 0.74:0.78; Spanish: 0.70, CI = 0.66:0.74; English: 0.79, CI = 0.76:0.82

**Table 4.** Optimal MoCA cutoffs to distinguish between MCI and dementia.

| Table 4. Optimal MoCA cutoffs to distinguish between MCI and dementia |  |  |  |  |  |  |  |  |  |
| --- | --- | --- | --- | --- | --- | --- | --- | --- | --- |
| Score | Sensitivity |  |  | Specificity |  |  | Youden's Index |  |  |
|  | Total | Spanish | English | Total | Spanish | English | Total | Spanish | English |
| 15 | 0.67 | 0.76 | 0.53 | 0.89 | 0.79 | 0.99 | 0.56 | 0.55 | 0.52 |
| 16 | 0.71 | 0.80 | 0.59 | 0.86 | 0.76 | 0.98 | **0.57** | **0.56** | 0.57 |
| 17 | 0.76 | 0.82 | 0.68 | 0.81 | 0.68 | 0.95 | **0.57** | 0.5 | **0.63** |
| 18 | 0.82 | 0.87 | 0.74 | 0.74 | 0.61 | 0.88 | 0.56 | 0.48 | 0.62 |
| 19 | 0.87 | 0.91 | 0.82 | 0.67 | 0.54 | 0.80 | 0.54 | 0.45 | 0.62 |
| 20 | 0.89 | 0.94 | 0.82 | 0.61 | 0.48 | 0.75 | 0.5 | 0.42 | 0.57 |
| 21 | 0.92 | 0.96 | 0.87 | 0.53 | 0.41 | 0.66 | 0.45 | 0.37 | 0.53 |
| AUC: Total = 0.86. CI = 0.84:0.89; Spanish: 0.85. CI = 0.82:0.88; English: 0.89. CI = 0.86:0.92 |  |  |  |  |  |  |  |  |  |

Optimal cutoff scores for the MoCA Total Raw Score for detecting MCI or dementia stratified by age and education separately are presented in Supplemental tables 1 to 4. Cutoff scores decreased with older age and fewer years of education in both languages, with consistently lower cutoff scores among participants tested in Spanish. Tables 5 and 6 show the optimal cutoffs of the MoCA Total Raw Score to detect MCI or dementia by age and education combined. The pattern of lower cutoffs at older ages and lower educational attainment persisted across both languages. For example, a Spanish-tested individual with 6 or fewer years of education and 75 or older had a cutoff score of 9 for dementia but would have a cutoff of 19 with 16 or more years of education and under 75. If the second individual (16+ education and under 75) was an English speaker, their cutoff would be 20. Based on these stratified results, we provide an Excel based calculator to determine the optimal MoCA cutoff score based on language, age, and educational attainment (Supplemental Material 1).

**Table 5.** Optimal MoCA cutoffs to distinguish between normal cognition and MCI by language and a combined age/education variable among language-matched participants.

|  | Age Group | Education Group | Total Positive | Total Negative | Cutoff | AUC | Lower 95%CI | Upper 95%CI | Sen | Specificity |
| --- | --- | --- | --- | --- | --- | --- | --- | --- | --- | --- |
| Spanish | All | All | 241 | 342 | 22.1 | 0.7 | 0.66 | 0.73 | 0.76 | 0.54 |
|  | <=75yrs | <=6 | 28 | 44 | 17.4 | 0.67 | 0.55 | 0.77 | 0.61 | 0.68 |
|  |  | 7-11 | 23 | 45 | 19.5 | 0.82 | 0.73 | 0.90 | 0.7 | 0.71 |
|  |  | 12-15 | 56 | 80 | 23 | 0.75 | 0.68 | 0.82 | 0.82 | 0.59 |
|  |  | 16+ | 36 | 75 | 23.8 | 0.76 | 0.68 | 0.83 | 0.53 | 0.8 |
|  | >75yrs | <=6 | 19 | 35 | 14.2 | 0.77 | 0.65 | 0.89 | 0.63 | 0.77 |
|  |  | 7-11 | 14 | 22 | 17.9 | 0.72 | 0.56 | 0.86 | 0.64 | 0.73 |
|  |  | 12-15 | 26 | 23 | 20.2 | 0.8 | 0.69 | 0.89 | 0.62 | 0.87 |
|  |  | 16+ | 30 | 12 | 21.4 | 0.6 | 0.43 | 0.75 | 0.47 | 0.67 |
|  | Age Group | Education Group | Total Positive | Total Negative | Cutoff | AUC | Lower 95%CI | Upper 95%CI | Sen | Specificity |
| English | All | All | 557 | 220 | 24.2 | 0.79 | 0.76 | 0.82 | 0.8 | 0.66 |
|  | <=75yrs | NA | NA | NA | NA | NA |  |  | NA | NA |
|  |  | 7-11 | 7 | 7 | 22.2 | 0.76 | 0.51 | 0.97 | 1 | 0.43 |
|  |  | 12-15 | 63 | 180 | 24 | 0.72 | 0.66 | 0.70 | 0.63 | 0.65 |
|  |  | 16+ | 75 | 261 | 25.1 | 0.82 | 0.78 | 0.86 | 0.81 | 0.69 |
|  | >75yrs | NA | NA | NA | NA | NA | NA | NA | NA | NA |
|  |  | 7-11 | 8 | 5 | 13.6 | 0.51 | 0.21 | 0.79 | 1 | 0.2 |
|  |  | 12-15 | 40 | 57 | 23 | 0.76 | 0.67 | 0.83 | 0.82 | 0.58 |
|  |  | 16+ | 25 | 44 | 23.4 | 0.82 | 0.72 | 0.91 | 0.76 | 0.8 |

**Table 6.** Optimal MoCA cutoffs to distinguish between MCI and dementia by language and a combined age/education variable among language-matched participants.

|  | Age Group | Education Group | Total Positive | Total Negative | Cutoff | AUC | Lower 95%CI | Upper 95%CI | Sen | Specificity |
| --- | --- | --- | --- | --- | --- | --- | --- | --- | --- | --- |
| Spanish | All | All | 182 | 241 | 14.8 | 0.85 | 0.82 | 0.88 | 0.76 | 0.79 |
|  | <=75yrs | <=6 | 25 | 28 | 13.7 | 0.77 | 0.66 | 0.87 | 0.72 | 0.75 |
|  |  | 7-11 | 14 | 23 | 14.6 | 0.88 | 0.78 | 0.96 | 0.86 | 0.78 |
|  |  | 12-15 | 20 | 56 | 15.8 | 0.93 | 0.87 | 0.97 | 0.85 | 0.89 |
|  |  | 16+ | 18 | 36 | 18.7 | 0.83 | 0.72 | 0.93 | 0.78 | 0.78 |
|  | >75yrs | <=6 | 39 | 19 | 9.3 | 0.86 | 0.77 | 0.93 | 0.72 | 0.95 |
|  |  | 7-11 | 23 | 14 | 12.1 | 0.8 | 0.67 | 0.91 | 0.61 | 0.93 |
|  |  | 12-15 | 21 | 26 | 15.9 | 0.83 | 0.72 | 0.92 | 0.76 | 0.77 |
|  |  | 16+ | 15 | 30 | 18 | 0.77 | 0.63 | 0.90 | 0.6 | 0.7 |
|  | Age Group | Education Group | Total Positive | Total Negative | Cutoff | AUC | Lower 95%CI | Upper 95%CI | Sen | Specificity |
| English | All | All | 131 | 220 | 17.1 | 0.89 | 0.86 | 0.90 | 0.74 | 0.88 |
|  | <=75yrs | NA | NA | NA | NA | NA | NA | NA | NA | NA |
|  |  | 7-11 | 4 | 7 | 15 | 0.79 | 0.46 | 1.0 | 0.5 | 1 |
|  |  | 12-15 | 43 | 63 | 17.5 | 0.93 | 0.89 | 0.97 | 0.79 | 0.92 |
|  |  | 16+ | 32 | 75 | 19.9 | 0.86 | 0.78 | 0.93 | 0.69 | 0.83 |
|  | >75yrs | NA | NA | NA | NA | NA | NA | NA | NA | NA |
|  |  | 7-11 | 7 | 8 | 16.8 | 0.87 | 0.70 | 0.99 | 0.71 | 0.75 |
|  |  | 12-15 | 18 | 40 | 16 | 0.93 | 0.85 | 0.99 | 0.72 | 1 |
|  |  | 16+ | 24 | 25 | 17.8 | 0.88 | 0.79 | 0.95 | 0.67 | 0.92 |

## Discussion

In this secondary cross-sectional data analysis of the NACC UDS 3.0, we aimed to identify optimal MoCA cutoff scores for MCI and dementia among US Latinos. This work addresses key limitations of prior studies by using a large sample, separating MCI and dementia as reference standards, restricting analyses to participants tested in their primary language, and stratifying results by language, education, and age. Consistent with our hypothesis, Spanish-tested participants, older adults, and individuals with lower educational attainment required lower cutoff scores to optimally detect MCI and dementia.

In contrast to the MoCA cutoff score for designating cognitive impairment of 26 [22], our findings indicate that lower cutoff scores are more appropriate for US Latinos. Specifically, the optimal cutoff for MCI was 23 among participants tested in Spanish and 25 for those tested in English. The standard one-point education correction was insufficient to account for educational differences. Among English-tested participants with 7 to 11 years of education, the optimal cutoff was 23 [22], which remained several points below the established threshold even after correction. Differences were more pronounced among Spanish tested participants for whom the optimal cutoff scores were 15 for those with less than 6 years of education and 18.5 for those with 7 to 11 years of education. These values are nearly 10 points below the established cutoff, even after applying the standard education adjustment. Prior examinations of the NACC data did not include participants with fewer than 7 years of education [19], underscoring the limitations of existing MoCA scoring approaches for identifying MCI, particularly among Spanish tested individuals with low educational attainment.

Age also exerted an independent influence on optimal cutoff scores. Across educational strata, participants aged 75 years or older required cutoff scores that were approximately 2 to 3 points lower than those of younger participants. These findings indicate that any adjustment to MoCA raw scores must jointly consider age, educational attainment, and language of testing. Importantly, all participants in the present analyses were tested in their primary language. As a result, the lower cutoffs identified in this study cannot be attributed to reduced proficiency in the language of test administration.

Several limitations warrant consideration. The NACC UDS is not a probability-based population sample, which limits generalizability of findings. However, the dataset includes substantial heterogeneity in age, educational attainment, and country of birth among Latino participants [28]. Although this study included a larger Latino sample than prior investigations, sample size constraints limited finer stratification, particularly among English tested participants with very low educational attainment. The diagnostic tools utilized during the consensus process include the MoCA among other neuropsychological tests, which may have introduced bias and potentially inflated estimates of diagnostic accuracy.

Despite these limitations, these findings have important implications for both clinical practice and research. This study provides the most detailed estimates to date of optimal MoCA cutoff scores for US Latinos accounting for language, age, and education. Use of these stratified cutoffs may reduce misclassification during cognitive screening, improve diagnostic accuracy, and enhance equity in dementia care. Furthermore, the MoCA is widely used in research for eligibility or outcome purposes. These findings may improve eligibility determination and outcome classification for Latino participants in observational and interventional studies.

Future studies should expand sample sizes to allow finer stratification across demographic factors and should evaluate optimal cutoff scores in probability-based samples of US Latinos. Longitudinal studies are also needed to assess predictive validity, including associations with cognitive decline trajectories and neuroimaging biomarkers. Additional work may examine whether combining the MoCA with other screening tools adds incremental validity and improves diagnostic accuracy.

Accurate cognitive screening tools are essential for identifying MCI and dementia among US Latinos. As the Latino population ages and research increases, reliance on the standard MoCA scoring procedure risks continued misdiagnosis in clinical settings and misclassification in research. Our study demonstrates that the MoCA can be used effectively among US Latinos when cutoff scores account for language, age and educational attainment. Continued investigation is needed to strengthen the evidence base and refine screening approaches for this growing and underserved population.

## Data Availability

All data produced in the present study are available upon reasonable request to the authors.

## Acknowledgements

The NACC database is funded by NIA/NIH Grant U24 AG072122. NACC data are contributed by the NIA-funded ADRCs: P30 AG062429 (PI James Brewer, MD, PhD), P30 AG066468 (PI Oscar Lopez, MD), P30 AG062421 (PI Bradley Hyman, MD, PhD), P30 AG066509 (PI Thomas Grabowski, MD), P30 AG066514 (PI Mary Sano, PhD), P30 AG066530 (PI Helena Chui, MD), P30 AG066507 (PI Marilyn Albert, PhD), P30 AG066444 (PI David Holtzman, MD), P30 AG066518 (PI Lisa Silbert, MD, MCR), P30 AG066512 (PI Thomas Wisniewski, MD), P30 AG066462 (PI Scott Small, MD), P30 AG072979 (PI David Wolk, MD), P30 AG072972 (PI Charles DeCarli, MD), P30 AG072976 (PI Andrew Saykin, PsyD), P30 AG072975 (PI Julie A. Schneider, MD, MS), P30 AG072978 (PI Ann McKee, MD), P30 AG072977 (PI Robert Vassar, PhD), P30 AG066519 (PI Frank LaFerla, PhD), P30 AG062677 (PI Ronald Petersen, MD, PhD), P30 AG079280 (PI Jessica Langbaum, PhD), P30 AG062422 (PI Gil Rabinovici, MD), P30 AG066511 (PI Allan Levey, MD, PhD), P30 AG072946 (PI Linda Van Eldik, PhD), P30 AG062715 (PI Sanjay Asthana, MD, FRCP), P30 AG072973 (PI Russell Swerdlow, MD), P30 AG066506 (PI Glenn Smith, PhD, ABPP), P30 AG066508 (PI Stephen Strittmatter, MD, PhD), P30 AG066515 (PI Victor Henderson, MD, MS), P30 AG072947 (PI Suzanne Craft, PhD), P30 AG072931 (PI Henry Paulson, MD, PhD), P30 AG066546 (PI Sudha Seshadri, MD), P30 AG086401 (PI Erik Roberson, MD, PhD), P30 AG086404 (PI Gary Rosenberg, MD), P20 AG068082 (PI Angela Jefferson, PhD), P30 AG072958 (PI Heather Whitson, MD), P30 AG072959 (PI James Leverenz, MD).

## Statements and Declarations; Compliance with Ethical Standards

- Disclosure of potential conflicts of interest: The authors have no relevant financial or non-financial interests to disclose.
- Research involving Human Participants and/or Animals: All ADRCs received approval to gather data at their institutional review board.
- Informed consent: Written informed consent and/or assent was obtained from all participants at the individual ADRCs.

## Data availability statement

De-identified data is available from NACC at the following website: https://naccdata.org/

---

**Supplemental table 1.** Optimal MoCA cutoffs to distinguish between normal cognition and MCI by language and age (<=75yrs vs >75) among language-matched participants.

|  | Age Group | Total Positive | Total Negative | Cutoff | AUC | Lower 95%CI | Upper 95%CI | Sen | Specificity |
| --- | --- | --- | --- | --- | --- | --- | --- | --- | --- |
| Spanish | $\leq 75$ yrs | 146 | 250 | 22.5 | 0.73 | 0.69 | 0.77 | 0.74 | 0.62 |
| | $> 75$ yrs | 95 | 92 | 16.8 | 0.61 | 0.54 | 0.68 | 0.37 | 0.77 |
| English | $\leq 75$ yrs | 145 | 451 | 24.7 | 0.78 | 0.74 | 0.81 | 0.75 | 0.69 |
| | $> 75$ yrs | 75 | 106 | 23.1 | 0.78 | 0.72 | 0.84 | 0.83 | 0.65 |

**Supplemental table 2.** Optimal MoCA cutoffs to distinguish between normal cognition and MCI by language and education (<=6 vs 7-11 vs 12-15 vs 16+) among language-matched participants.

|  | Education Group | Total Positive | Total Negative | Cutoff | AUC | Lower 95%CI | Upper 95%CI | Sen | Specificity |
| --- | --- | --- | --- | --- | --- | --- | --- | --- | --- |
| Spanish | $\leq 6$ | 47 | 79 | 15.1 | 0.70 | 0.61 | 0.78 | 0.53 | 0.77 |
|  | 7-11 | 37 | 67 | 18.5 | 0.78 | 0.71 | 0.82 | 0.65 | 0.81 |
|  | 12-15 | 82 | 103 | 22.4 | 0.76 | 0.74 | 0.82 | 0.74 | 0.65 |
|  | 16+ | 66 | 87 | 23.5 | 0.75 | 0.68 | 0.81 | 0.58 | 0.76 |
| English | <=6 | NA | NA | NA | NA | NA | NA | NA | NA |
|  | 7-11 | 15 | 12 | 22.5 | 0.66 | 0.47 | 0.84 | 0.93 | 0.33 |
|  | 12-15 | 103 | 237 | 23.4 | 0.74 | 0.69 | 0.78 | 0.71 | 0.63 |
|  | 16+ | 100 | 305 | 24.7 | 0.82 | 0.78 | 0.86 | 0.71 | 0.78 |

**Supplemental table 3.** Optimal MoCA cutoffs to distinguish between MCI and dementia by language and age (<=75yrs vs >75) among language-matched participants.

|  | Age Group | Total Positive | Total Negative | Cutoff | AUC | Lower 95%CI | Upper 95%CI | Sen | Specificity |
| --- | --- | --- | --- | --- | --- | --- | --- | --- | --- |
| Spanish | <=75yrs | 78 | 146 | 15.3 | 0.86 | 0.81 | 0.90 | 0.77 | 0.80 |
|  | >75yrs | 104 | 95 | 13.5 | 0.84 | 0.79 | 0.88 | 0.70 | 0.79 |
| English | <=75yrs | 80 | 145 | 17.7 | 0.89 | 0.85 | 0.93 | 0.71 | 0.90 |
|  | >75yrs | 51 | 75 | 16.6 | 0.89 | 0.83 | 0.94 | 0.71 | 0.95 |

**Supplemental table 4.** Optimal MoCA cutoffs to distinguish between MCI and dementia by language and education (<=6 vs 7-11 vs 12-15 vs 16+) among language-matched participants.

|  | Education Group | Total Positive | Total Negative | Cutoff | AUC | Lower 95%CI | Upper 95%CI | Sen | Specificity |
| --- | --- | --- | --- | --- | --- | --- | --- | --- | --- |
| Spanish | <=6 | 64 | 47 | 11.5 | 0.83 | 0.76 | 0.88 | 0.70 | 0.74 |
|  | 7-11 | 37 | 37 | 13.1 | 0.84 | 0.76 | 0.91 | 0.70 | 0.78 |
|  | 12-15 | 41 | 82 | 16.0 | 0.89 | 0.84 | 0.94 | 0.80 | 0.85 |
|  | 16+ | 33 | 66 | 18.1 | 0.81 | 0.72 | 0.88 | 0.70 | 0.74 |
| English | <=6 | NA | NA | NA | NA | NA | NA | NA | NA |
|  | 7-11 | 11 | 15 | 15.9 | 0.83 | 0.69 | 0.96 | 0.55 | 0.87 |
|  | 12-15 | 61 | 103 | 16.4 | 0.92 | 0.89 | 0.96 | 0.75 | 0.95 |
|  | 16+ | 56 | 100 | 18.8 | 0.87 | 0.82 | 0.92 | 0.73 | 0.86 |

## Reference List

1. Pan American Health Organization, Leading causes of mortality and health loss at the regional, subregional, and country levels in the Region of the Americas, 2000-2019. 2021: https://www.paho.org/en/noncommunicable-diseases-and-mental-health/noncommunicable-diseases-and-mental-health-data-39.

2. Hurd, M.D., et al., Monetary costs of dementia in the United States. New England Journal of Medicine, 2013. 368(14): p. 1326–1334.

3. National Institute on Aging. Goal F: Understand health disparities and develop strategies to improve the health status of older adults in diverse populations 2018.

4. U.S. Department of Health & Human Services, National Plan to Address Alzheimer’s Disease: 2022 update, O.o.T.A.S.f.P.a. Evaluation, Editor. 2022.

5. Hudomiet, P., M.D. Hurd, and S. Rohwedder, Trends in inequalities in the prevalence of dementia in the United States. Proceedings of the National Academy of Sciences, 2022. 119(46): p. e2212205119.

6. Wu, S., et al., Latinos & Alzheimer’s Disease: New numbers behind the crisis. Projection of the costs for U.S. Latinos living with Alzheimer’s Disease through 2060. 2016, USC Edward R. Roybal Institute on Aging and the LatinosAgainstAlzheimer’s Network.

7. Lin, P.J., et al., Racial and Ethnic Differences in Knowledge About One’s Dementia Status. Journal of the American Geriatrics Society, 2020. 68(8): p. 1763–1770.

8. Barker, W.W., et al., The effect of a memory screening program on the early diagnosis of Alzheimer disease. Alzheimer Disease & Associated Disorders, 2005. 19(1): p. 1–7.

9. Hinton, L., et al., Mapping racial and ethnic healthcare disparities for persons living with dementia: A scoping review. Alzheimer’s & Dementia, 2024. 20(4): p. 3000–3020.

10. Cotter, V.T., Alzheimer’s disease: issues and challenges in primary care. Nursing Clinics, 2006. 41(1): p. 83–93.

11. Gitlin, L.N., H.C. Kales, and C.G. Lyketsos, Nonpharmacologic management of behavioral symptoms in dementia. Jama, 2012. 308(19): p. 2020–2029.

12. Hinton, L., et al., Dementia neuropsychiatric symptom severity, help-seeking patterns, and family caregiver unmet needs in the Sacramento Area Latino Study on Aging (SALSA). Clinical gerontologist, 2006. 29(4): p. 1–15.

13. Salazar, R., A.K. Dwivedi, and D.R. Royall, Cross-ethnic differences in the severity of neuropsychiatric symptoms in persons with mild cognitive impairment and Alzheimer’s disease. The Journal of neuropsychiatry and clinical neurosciences, 2017. 29(1): p. 13–21.

14. Chen, C., J. Thunell, and J. Zissimopoulos, Changes in physical and mental health of Black, Hispanic, and White caregivers and non-caregivers associated with onset of spousal dementia. Alzheimer’s & Dementia: Translational Research & Clinical Interventions, 2020. 6(1): p. e12082.

15. Galvin, J.E. and C.H. Sadowsky, Practical guidelines for the recognition and diagnosis of dementia. The Journal of the American Board of Family Medicine, 2012. 25(3): p. 367–382.

16. Garcia, D.M., M.M. Pinzon, and J. Perales-Puchalt, Understanding Healthcare Barriers for Latino/a/e/x Families with Alzheimer’s Disease: Insights from Primary Care Provider interviews. medRxiv, 2024.

17. Mungas, D., et al., Age and education correction of Mini-Mental State Examination for English-and Spanish-speaking elderly. Neurology, 1996. 46(3): p. 700–706.

18. Zhou, Y., et al., Use of the MoCA in detecting early Alzheimer’s disease in a Spanish-speaking population with varied levels of education. Dementia and geriatric cognitive disorders extra, 2015. 5(1): p. 85–95.

19. Milani, S.A., et al., Optimal cutoffs for the Montreal Cognitive Assessment vary by race and ethnicity. Alzheimer’s & Dementia: Diagnosis, Assessment & Disease Monitoring, 2018. 10: p. 773–781.

20. Stimmel, M.B., et al., Is the Montreal cognitive assessment culturally valid in a diverse geriatric primary care setting? Lessons from the Bronx. Journal of the American Geriatrics Society, 2024. 72(3): p. 850–857.

21. Manly, J.J., et al., Telephone-based identification of mild cognitive impairment and dementia in a multicultural cohort. Archives of neurology, 2011. 68(5): p. 607–614.

22. Nasreddine, Z.S., et al., The Montreal Cognitive Assessment, MoCA: a brief screening tool for mild cognitive impairment. Journal of the American Geriatrics Society, 2005. 53(4): p. 695–699.

23. Marquine, M.J., et al., Demographically-adjusted normative data among Latinos for the version 3 of the Alzheimer’s Disease Centers’ Neuropsychological Test Battery in the Uniform Data Set. Alzheimer’s & Dementia, 2023.

24. Beekly, D.L., et al., The National Alzheimer’s Coordinating Center (NACC) database: the uniform data set. Alzheimer Disease & Associated Disorders, 2007. 21(3): p. 249–258.

25. Weintraub, S., et al., The Alzheimer’s Disease Centers’ Uniform Data Set (UDS): the neuropsychologic test battery. Alzheimer Dis Assoc Disord, 2009. 23(2): p. 91–101.

26. Robin, X., et al., pROC: an open-source package for R and S+ to analyze and compare ROC curves. BMC bioinformatics, 2011. 12(1): p. 77.

27. Thiele, C. and G. Hirschfeld, cutpointr: improved estimation and validation of optimal cutpoints in R. Journal of Statistical Software, 2021. 98: p. 1–27.

28. Perales-Puchalt, J., et al., Risk of mild cognitive impairment among older adults in the United States by ethnoracial group. International psychogeriatrics, 2021. 33(1): p. 51–62.

